# Development of Comorbidity Index for In-hospital Mortality for Patients Underwent Coronary Artery Revascularization

**DOI:** 10.1101/2023.04.08.23288311

**Authors:** Renxi Li

**Author notes:** **Corresponding Author:** Renxi Li, 2300 I St NW, Washington, DC 20052. The George Washington University School of Medicine and Health Sciences, Department of Surgery. **Informed Patient Consent:** This study excepted from IRB approval because it used retrospective, de-identified NIS data.

## Abstract

**Background:** For myocardial revascularization, coronary artery bypass grafting (CAGB) and percutaneous coronary intervention (PCI) are two common modalities but with high in-hospital mortality. A comorbidity index is useful to predict mortality or can be used with other covariates to develop point-scoring systems. This study aimed to develop specific comorbidity indices for patients who underwent coronary artery revascularization.

**Methods:** Patients who underwent CABG or PCI were identified in the National Inpatient Sample database between Q4 2015-2020. Patients of age<40 were excluded for congenital heart defects. Patients were randomly sampled into experimental (70%) and validation (30%) groups. Thirty-eight Elixhauser comorbidities were identified and included in multivariable regression to predict in-hospital mortality. Weight for each comorbidity was assigned and single indices, Li CABG Mortality Index (LCMI) and Li PCI Mortality Index (LPMI), were developed.

**Results:** Mortality prediction by LCMI approached adequacy (*c*-statistic=0.691, 95% CI=0.682-0.701) and was comparable to multivariable regression with comorbidities (*c*-statistic=0.685, 95% CI=0.675-0.694). LCMI prediction performed significantly better than Elixhauser Comorbidity Index (ECI) (*c*-statistic=0.621, 95% CI=0.611-0.631) and can be further improved by adjusting age (*c*-statistic=0.721, 95% CI=0.712-0.730).

LPMI moderately predicted in-hospital mortality (*c*-statistic=0.666, 95% CI=0.660-0.672) and performed significantly better than ECI (*c*-statistic=0.610, 95% CI=0.604-0.616). LPMI performed better than the all-comorbidity multivariable regression (*c*-statistic=0.658, 95% CI=0.652-0.663). After age adjustment, LPMI prediction was significantly increased and was approaching adequacy (*c*-statistic=0.695, 95% CI=0.690-0.701).

**Conclusions:** LCMI and LPMI effectively predicted in-hospital mortality. These indices were validated and performed superior to ECI. The adjustment of age increased their predictive power to adequacy, implicating potential clinical application.

## Introduction

Coronary artery disease (CAD) is a prevalent cause of mortality that accounts for 1 in 4 death in the United States ^1^. For CAD patients indicated for myocardial revascularization, coronary artery bypass grafting (CAGB) surgery and percutaneous coronary intervention (PCI) are the two most common modalities ^2^. CABG and PCI are high-risk procedures with crude in-hospital mortality rates between 2-4 percent and 1-2 percent, respectively ^3–11^.

When assessing clinical outcomes and considering the associated risks, it is often necessary to factor in comorbidities ^12,13^. There are many comorbidity measures, of which Elixhauser Comorbidity was an established modality for identifying comorbidities in large administrative datasets ^14^. Comorbidities can be used to predict the likelihood of in-hospital mortality and numerous studies have shown the efficacy of the Elixhauser measures in this application ^15–17^. Elixhauser Comorbidity measure was developed into the Elixhauser Comorbidity Index (ECI), which is a single index that predicts the likelihood of in-hospital mortality ^18^. However, ECI was developed in all diseases and the validation of ECI showed decreased efficacy in some specific disease categories ^18^. Thus, ECI might be overly general; CABG- and PCI-specific mortality indices would be more useful to discriminate in-hospital mortality in patients who underwent these procedures.

This study aimed to develop specific comorbidity indices for patients who underwent coronary artery revascularization using the National (Nationwide) Inpatient Sample (NIS) database, the largest in-patient database in the United States ^19^.

## Methods

The NIS database was utilized to gather data on patients who underwent CABG and PCI between the final quarter of 2015 and 2020. All cases were collected using the International Classification of Diseases, Tenth Revision, Procedure Coding System (ICD-10-PCS) codes: 0210xxxx for CABG; 02703xx, 02713xx, 02723xx, and 02733xx for PCI (027×3Tx and 027×3Zx excluded). Patients of age younger than 40 years old were excluded for congenital heart defects. Patient age, in-hospital mortality rates, and International Classification of Diseases, Tenth Revision, Clinical Modification (ICD-10-CM) codes were extracted. Furthermore, the Elixhauser Comorbidity Software was employed to identify the presence of thirty-eight different comorbidities ^20^.

In the CABG and PCI groups, respectively, patients were randomly sampled into an experimental group (70% of all cases) to develop the mortality indices. The rest of the patients (30% of all cases) were assigned as the validation group to verify the indices. In the CABG and PCI experimental groups, respectively, all thirty-eight comorbidities were included in the multivariable logistic regression to predict in-hospital mortality. These comorbidities were then developed into single indices, Li CABG Mortality Index (LCMI) and Li PCI Mortality Index (LPMI), for CABG and PCI groups, respectively, according to Sullivan’s method ^10,21^. The parameter estimates of each comorbidity from the multivariable logistic regression were extracted. Weight for each comorbidity was calculated by dividing its regression coefficient by the absolute value of the smallest regression coefficient, followed by rounding the quotient to the nearest integer value. Comorbidity with a larger coefficient thus signified more contribution to the mortality index respective to the comorbidity with the smallest coefficient.

The predictive power of LCMI and LPMI were examined in their respective validation groups. Moreover, the predictive powers of ECI were examined in the experimental groups and then compared to those of LCMI and LPMI. To show the practicality of LCMI and LPMI, age, as an established risk factor for mortality in coronary revascularization, was included in the multivariable logistic regression of the new mortality indices to predict in-hospital mortality.

A graph was created for the receiver operating characteristic (ROC) of each regression. The area under the ROC curve (AUC) was calculated, and a 95% confidence interval was determined. The ability of each model to distinguish in-hospital mortality was assessed using the *c*-statistic, which ranges from 0.5 (no predictive power) to 1 (accurate classification of all in-hospital deaths). A *c*-statistic greater than 0.7 indicates the presence of adequate predictive power. A *c*-statistic between 0.6 and 0.7 denotes moderate predictive power. Comparisons of two different AUCs were conducted using the DeLong test ^22^.

All analyses were performed by SAS (version 9.4). The study was retrospective and was based on the open database and thus was IRB-exempted. The author had full access to all data and takes responsibility for the integrity of data analysis.

## Results

There were 189,662 patients who underwent CABG identified in NIS between the final quarter of 2015 and 2020. There were 133,378 cases sampled into the experimental group and the rest 56,284 cases were assigned to the validation group. For patients who underwent PCI, there were 431,021 cases identified. A total number of 304,888 cases were sampled into the experimental group and the rest 126,133 cases were assigned to the validation group.

The comorbidities and the weights of each comorbidity to derive LCMI and LPMI in CABG and PCI experimental groups, respectively, were summarized in Table 1. Comorbidities that were not presented were assigned a weight of 0.

**Table 1.** Comorbidities and their respective weights to derive LCMI and LPMI in CABG and PCI experimental groups. Patients were identified in the NIS database from the last quarter of 2015 to 2020. ***Abbreviations:*** CABG, Coronary Artery Bypass Surgery; LCMI, Li CABG Mortality Index; LPMI, Li PCI Mortality Index; NIS, National (Nationwide) Inpatient Sample; PCI, Percutaneous Coronary Intervention.

| Comorbidities | CABG, n (%) | CABG Weight | PCI, n (%) | PCI Weight |
| --- | --- | --- | --- | --- |
| Acquired immune deficiency syndrome | 480 (0.36%) | 3 | 1305 (0.43%) | -22 |
| Alcohol abuse | 4413 (3.31%) | -4 | 8678 (2.85%) | 4 |
| Anemias due to other nutritional deficiencies | 0 (0.00%) | 0 | 0 (0.00%) | 0 |
| Autoimmune conditions | 3416 (2.56%) | -1 | 8201 (2.69%) | 5 |
| Chronic blood loss anemia (iron deficiency) | 0 (0.00%) | 0 | 0 (0.00%) | 0 |
| Lymphoma | 471 (0.35%) | 5 | 1317 (0.43%) | 36 |
| Leukemia | 574 (0.43%) | 8 | 1169 (0.38%) | 30 |
| Metastatic cancer | 307 (0.23%) | 14 | 1624 (0.53%) | 67 |
| Solid tumor without metastasis, in situ | 25 (0.02%) | 9 | 40 (0.01%) | 1 |
| Solid tumor without metastasis, malignant | 1759 (1.32%) | 3 | 4017 (1.32%) | 20 |
| Cerebrovascular disease | 2640 (1.98%) | -6 | 4996 (1.64%) | 11 |
| Heart failure | 105 (0.08%) | 16 | 110 (0.04%) | 63 |
| Coagulopathy | 0 (0.00%) | 0 | 0 (0.00%) | 0 |
| Dementia | 1493 (1.12%) | 6 | 7158 (2.35%) | 42 |
| Depression | 11639 (8.73%) | -7 | 25872 (8.49%) | 25 |
| Diabetes without chronic complications | 21928 (16.44%) | -5 | 55273 (18.15%) | 11 |
| Diabetes with chronic complications | 41914 (31.42%) | -6 | 68216 (22.40%) | -7 |
| Drug abuse | 2276 (1.71%) | -4 | 6668 (2.19%) | -13 |
| Hypertension, complicated | 48853 (36.63%) | -2 | 95876 (31.48%) | -2 |
| Hypertension, uncomplicated | 67646 (50.72%) | -20 | 147189 (48.33%) | -62 |
| Liver disease, mild | 0 (0.00%) | 0 | 0 (0.00%) | 0 |
| Liver disease, moderate to severe | 116 (0.09%) | 3 | 324 (0.11%) | -14 |
| Chronic pulmonary disease | 28906 (21.67%) | 4 | 58792 (19.30%) | 3 |
| Neurological disorders affecting movement | 0 (0.00%) | 0 | 0 (0.00%) | 0 |
| Other neurological disorders | 0 (0.00%) | 0 | 0 (0.00%) | 0 |
| Seizures and epilepsy | 0 (0.00%) | 0 | 0 (0.00%) | 0 |
| Obesity | 37880 (28.40%) | -4 | 64835 (21.29%) | -21 |
| Paralysis | 1901 (1.43%) | 14 | 3801 (1.25%) | 15 |
| Peripheral vascular disease | 20027 (15.02%) | 7 | 32170 (10.56%) | 28 |
| Psychoses | 0 (0.00%) | 0 | 0 (0.00%) | 0 |
| Pulmonary circulation disease | 0 (0.00%) | 0 | 0 (0.00%) | 0 |
| Renal failure, moderate | 0 (0.00%) | 0 | 0 (0.00%) | 0 |
| Renal failure, severe | 3116 (2.34%) | 10 | 9270 (3.04%) | 4 |
| Hypothyroidism | 14854 (11.14%) | -2 | 34280 (11.26%) | -5 |
| Other thyroid disorders | 1426 (1.07%) | -4 | 2405 (0.79%) | -28 |
| Peptic ulcer disease x bleeding | 0 (0.00%) | 0 | 0 (0.00%) | 0 |
| Valvular disease | 1638 (1.23%) | 12 | 4076 (1.34%) | -16 |
| Weight loss | 0 (0.00%) | 0 | 0 (0.00%) | 0 |

The summary and comparison of the AUCs of different CABG models were shown in Table 2 and Figure 1. In the CABG experimental group, the prediction of in-hospital mortality by LCMI was approaching adequacy (*c*-statistic = 0.691, 95% CI = 0.682-0.701). In the validation group of CABG, the predictive power of LCMI (*c*-statistic = 0.685, 95% CI = 0.671-0.699) was similar (DeLong test p = 0.373) to that in the experimental group. The single index LCMI had comparable (DeLong test p = 0.245) predictive power as the multivariable regression model including all dichotomous comorbidities (*c*-statistic = 0.685, 95% CI = 0.675-0.694). Compared to ECI (*c*-statistic = 0.621, 95% CI = 0.611-0.631), LCMI significantly better discriminated in-hospital mortality (DeLong test p <0.001). The prediction of in-hospital mortality by LCMI could be significantly improved by including age in the regression model (*c*-statistic = 0.721, 95% CI = 0.712-0.730, DeLong test p <0.001).

**Table 2.**
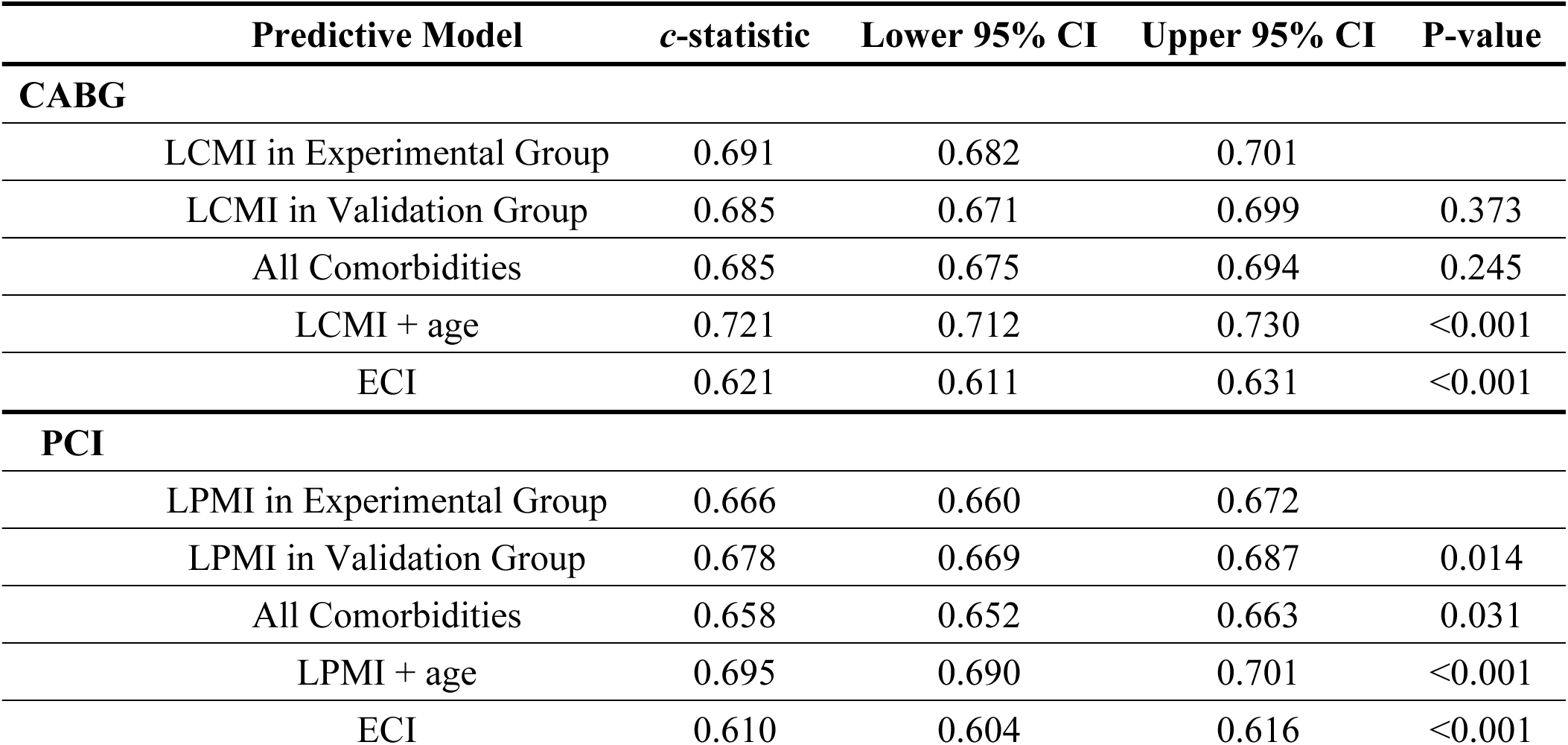
Assessment of in-hospital mortality discrimination by LCMI and LPMI in CABG and PCI groups, respectively. Patients were identified in the NIS database from the last quarter of 2015 to 2020. ***Abbreviations:*** CABG, Coronary Artery Bypass Surgery; CI, confidence interval; LCMI, Li CABG Mortality Index; LPMI, Li PCI Mortality Index; NIS, National (Nationwide) Inpatient Sample; PCI, Percutaneous Coronary Intervention.

**Figure 1.**
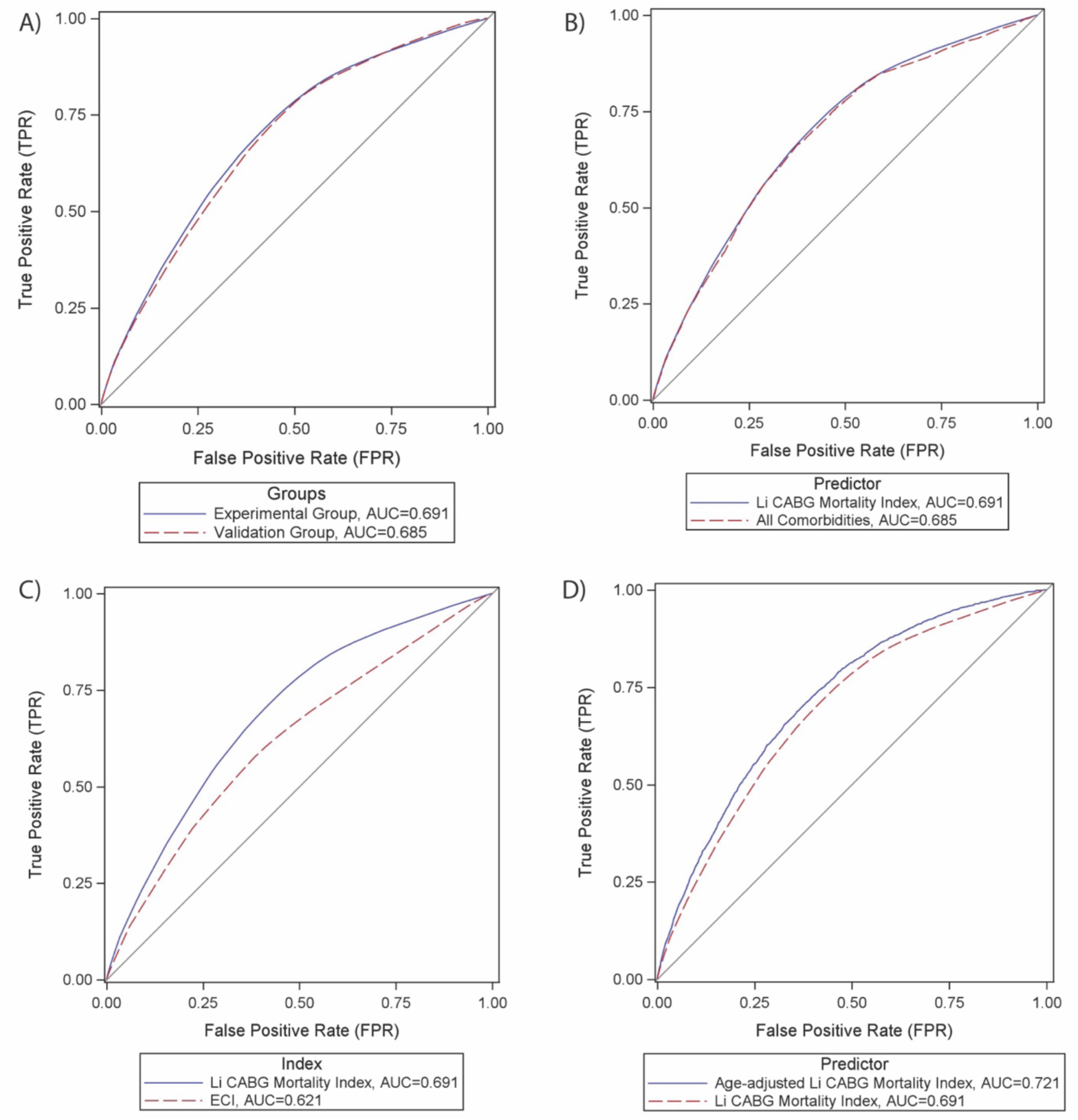
Comparison of ROC in predicting in-hospital mortality in patients who underwent CABG from the last quarter of 2015 to 2020. A) LCMI between experimental and the validation groups, B) between LCMI (experimental group) and multivariable regression model including all comorbidities, C) between LCMI (experimental group) and ECI, D) between LCMI and age-adjusted LCMI (experimental group). ***Abbreviations:*** CABG, Coronary Artery Bypass Surgery; LCMI, Li CABG Mortality Index; NIS, National (Nationwide) Inpatient Sample.

The AUCs of different PCI models were summarized and compared as shown in Table 2 and Figure 2. Although LPMI only moderately predicted in-hospital mortality in the experimental group of PCI (*c*-statistic = 0.666, 95% CI = 0.660-0.672), the predictive power of LPMI was significantly greater (DeLong test p <0.001) than that of ECI (*c*-statistic = 0.610, 95% CI = 0.604-0.616). Interestingly, the single index LPMI better discriminated (DeLong test p = 0.031) in-hospital mortality than the all-comorbidity multivariable regression (*c*-statistic = 0.658, 95% CI = 0.652-0.663). Also, the predictive power of LPMI (DeLong test p = 0.014) was higher in the validation group (*c*-statistic = 0.678, 95% CI = 0.669-0.687). Considering age into account, the prediction of in-hospital mortality by LPMI was significantly increased (DeLong test p <0.001) and was approaching adequacy (*c*-statistic = 0.695, 95% CI = 0.690-0.701).

**Figure 2.**
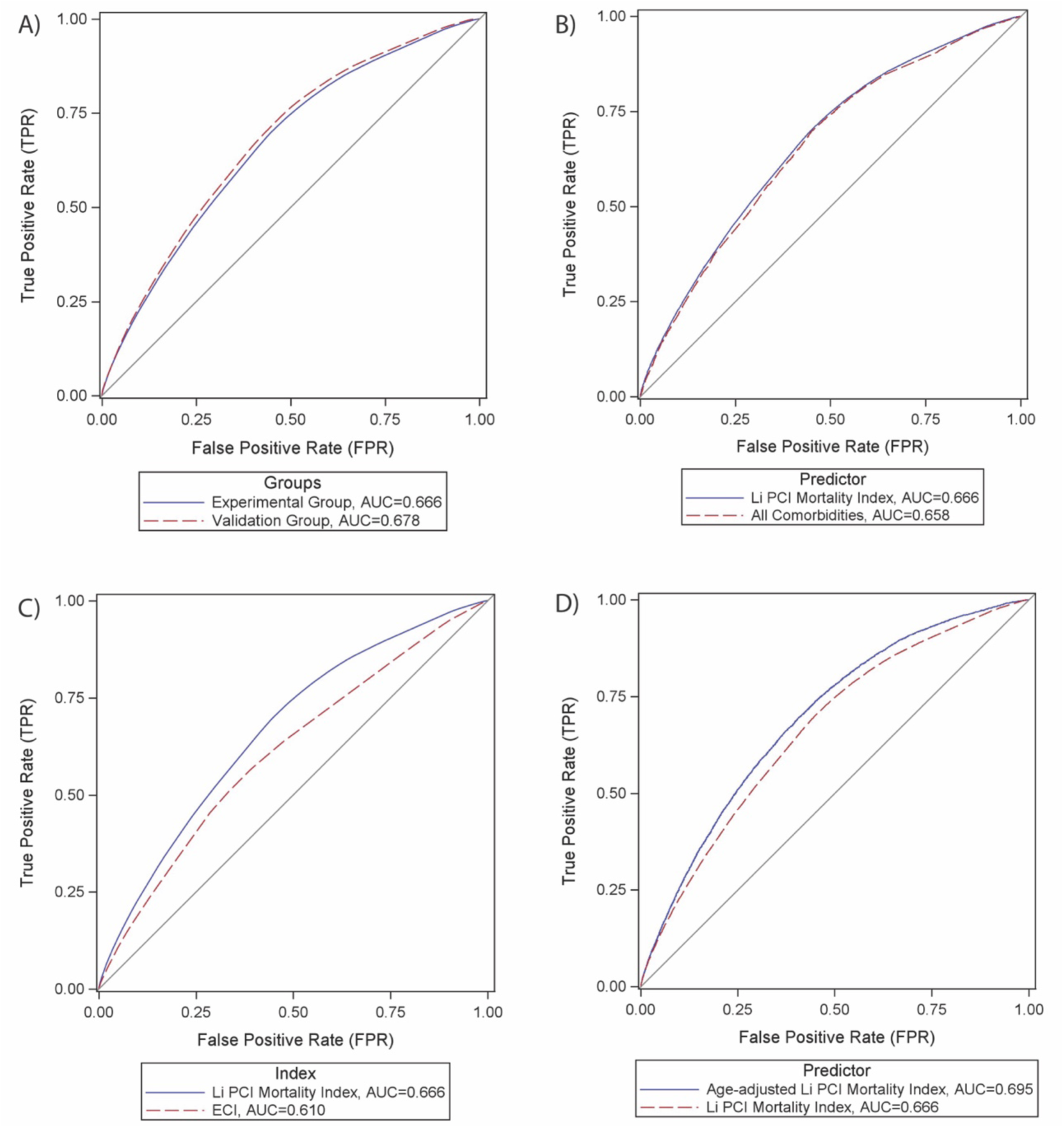
Comparison of ROC in predicting in-hospital mortality in patients who underwent PCI from the last quarter of 2015 to 2020. A) LPMI between experimental and the validation groups, B) between LPMI (experimental group) and multivariable regression model including all comorbidities, C) between LPMI (experimental group) and ECI, D) between LPMI and age-adjusted LCMI (experimental group). ***Abbreviations:*** LPMI, Li PCI Mortality Index; NIS, National (Nationwide) Inpatient Sample; PCI, Percutaneous Coronary Intervention.

## Discussion

LCMI was a well-developed index to predict in-hospital mortality in patients who underwent CABG. The predictive power of LCMI (*c*-statistic = 0.691) was approaching adequacy (*c*-statistic > 0.7). The AUC of LCMI did not differ (p = 0.245) from that of the multivariable regression with all dichotomous comorbidities, indicating LCMI well-translated information from all comorbidities into a single index. The AUC of LCMI did not differ (p = 0.373) between the experimental group and the validation group, indicating a good validity of the index. Future studies can further examine the validity of other data sources.

In patients who underwent PCI, LPMI moderately predicted (*c*-statistic = 0.666) in-hospital mortality. The AUC of LCMI was greater than (p = 0.031) the multivariable regression with comorbidities. Also, the AUC of LCMI between the experimental group and the validation group were different (p = 0.014). The AUCs of these three models were expected to match. In fact, the actual *c*-statistic did not differ to a great extent (experimental *c*-statistic = 0.666, multivariable comorbidities *c*-statistic = 0.658, validation *c*-statistic = 0.678). The contributor to a statistical difference might be due to a much larger sample size in PCI (PCI had 431,021 total cases while CABG had 189,662 cases), and thus a larger power that was more likely to signify differences in the PCI group.

Both LCMI and LPMI were significantly better predictors (p <0.001) for in-hospital mortality than ECI. This confirmed the hypothesis that ECI might be too general to use as an index for mortality prediction in CABG and PCI. It emphasized the necessity for developing specific indices for patients who underwent CABG and PCI and confirmed the significance of this study.

The LCMI and LPMI should be used in combination with other covariates. For example, in coronary artery revascularization, age is an established risk factor for mortality ^23–32^. After the development of the comorbidity indices, age was adjusted, and the accuracy of prediction was significantly improved to adequacy. Thus, despite the predictive power of LCMI and LPMI did not reach adequacy (*c*-statistic > 0.7), this showed the potential for them to be improved by including additional covariates. This way, the predictive power of mortality indices could be adequate enough to have real-life implications. Due to the limitations of the NIS database, some pertinent variables in coronary artery revascularization were not recorded. For future studies, relevant clinical, such as past medical history, specific diagnosis, lab values, as well as demographic information, can be added to adjust the LCMI and LPMI for more accurate assessments of in-hospital mortality risk. A new point-scoring system based on LCMI and/or LPMI and the covariates can be developed for easy assessment in the clinics.

There were several limitations of this study. First, the weights of the comorbidities in the indices were assigned based on mathematical weights in the regression. Additional assessment of the relative clinical significance of the comorbidities may be helpful to amend the indices. Second, the comorbidities were dichotomous so patients with varying degrees or stages of disease were assigned to same category. However, breaking down the comorbidities into more specific categories would decrease the statistical power and the usability of the indices. Nevertheless, physicians should be aware of this limitation when applying LCMI and LPMI.

In conclusion, LCMI and LPMI were developed in this study to predict in-hospital mortality CABG and PCI, respectively. These indices were validated and performed superior to ECI. The adjustment of age increased the predictive power of the LCMI and LPMI to adequacy, implicating potential application when including more clinically relevant covariates.

## Data Availability

All data produced in the present study are available upon reasonable request to the authors.

## Acknowledgements

Renxi Li participated in all aspects of this study. The author acknowledges the guidance and support of colleagues and reviewers who provided useful feedback throughout the work.

